# Knowledge, attitudes and behaviours related to tick awareness and prevention among users of urban green spaces: insights from stakeholder focus groups

**DOI:** 10.64898/2026.11.03.26366306

**Authors:** Jo Taylor-Egbeyemi, Sarah Masterton, Charlotte Robin

## Abstract

**Background:** Ticks in the UK can transmit pathogens that lead to diseases in humans, including Lyme disease and tick-borne encephalitis. Expanding tick populations and increasing human use of urban green spaces may elevate exposure risk. Public health advice exists, but little is known about how this guidance is understood and applied by users of urban environments.

**Methods:** Four stakeholder focus groups were conducted with professionals involved in tick fieldwork in urban green spaces. Discussions explored interactions with the public, perceived awareness, behaviours, and views on communication. Data were analysed thematically using an approach informed by the Framework Method.

**Results:** Stakeholders reported substantial variation in public awareness of ticks, influenced by prior exposure to rural environments and local knowledge. Misinformation, particularly around tick removal, was common. Signage was rarely observed in urban green spaces, and views on its effectiveness were mixed. Participants emphasised the need to balance risk communication with reassurance and highlighted the importance of age-appropriate education.

**Conclusions:** Insights from stakeholders underline the importance of targeted, context-specific public health messaging in urban settings. Priorities include identifying groups with lower awareness, addressing misinformation, designing balanced communication materials, evaluating the effectiveness of signage, and integrating education for children and families.

## Introduction

Ticks in the UK can carry pathogens that cause disease in humans, including Lyme disease [1]. With tick populations increasing in the UK [2], there is a risk of an increase in tick-borne infections.

Urban green spaces such as parks and gardens can provide suitable habitats for ticks. Activities such as walking on trails, sitting in grassy areas, or picnicking near bushes and long grass increase the likelihood of encountering ticks.

Dogs may also be vectors for ticks both into and between urban habitats [3]. As such, public health advice to reduce the risk of tick bites needs to account for *how* people use and interact with tick habitats, in addition to awareness of ticks and tick bite prevention. Public health advice in relation to the prevention and removal of tick bites is available, however it is not currently known how this advice is understood and enacted. Increased understanding of tick awareness and behaviour of people who use urban green spaces supports the development of public health campaigns to improve their efficacy [4].

Professionals who routinely interact with the public can provide valuable insight into public knowledge, behaviours, and misconceptions. Stakeholder perspectives can complement public-facing research and inform targeted public health interventions [5].

Therefore, the aim of the current study was to explore experiences of public engagement and perceived awareness and understanding of ticks with stakeholders who conduct fieldwork in urban green spaces. Gathering insights from stakeholders related to their experiences of interactions they have with members of the public during fieldwork will provide insight into perceived knowledge and behaviours relating to ticks.

## Methods

Four focus groups were conducted with UK Health Security Agency (UKHSA) staff and other stakeholders who have experience carrying out tick fieldwork, such as entomologists and surveillance staff. Each focus group contained between 2 and 4 participants, with 11 participants in total.

Participants were recruited via email through the UKHSA Medical Entomology and Zoonoses Ecology (MEZE) team. Eligible participants had prior experience conducting public-facing fieldwork activities, including tick surveillance and monitoring.

A topic guide was developed to explore:

- Participants’ interactions with members of the public
- Perceived public awareness of ticks and tick-borne disease
- Observed behaviours relevant to tick bite prevention in the public
- Views on public health messaging, signage, and educational needs

Groups were audio-recorded and, transcribed verbatim. Data were analysed using a reflexive thematic analysis approach informed by the Framework Method [5].

### Analysis

Transcripts were coded inductively and refined iteratively into a thematic framework.

### Ethics

The UKHSA Research Ethics and Governance Group (REGG) determined that formal ethical approval was not required as participants were participating in the project in their professional capacity.

## Results

Four themes and twelve subthemes were identified (Table S1).

### Public awareness, knowledge and preventive behaviours

Stakeholders reported substantial variation in public awareness of ticks and tick-borne infections. Stakeholders reported that individuals familiar with rural environments tended to possess higher awareness and adopt preventive behaviours such as tucking trousers into socks or checking for ticks after being outdoors. Other members of the public appeared surprised by the small size of ticks or held misconceptions regarding risk and removal techniques.

Local residents were reported to have strong practical knowledge of tick habitats and seasonal activity. Conversely, misinformation was widespread among other groups, including the use of Vaseline or twisting techniques to remove ticks. Participants also described inconsistencies in clinical advice from healthcare providers, noting that experiences with general practitioners varied considerably.

### Public perceptions and responses to tick risk

Stakeholders reported that many members of the public expressed curiosity when encountering field teams and engaged readily in conversations about ticks. Stakeholders reported that dog walkers frequently shared experiences of finding ticks on their pets. Emotional reactions varied: some individuals expressed fear or disgust, whereas others regarded ticks as an accepted aspect of outdoor life.

Stakeholders cautioned that risk communication must be carefully framed to avoid creating unnecessary concern. While heightened awareness is desirable, overly alarming messages may discourage outdoor activity or lead to unrealistic expectations of complete tick eradication.

### Communication and education

Signage relating to ticks was rarely observed in urban green spaces, and participants expressed mixed views about its usefulness. Some believed signage could effectively prompt preventive behaviours; others suggested it might induce worry or signal a more severe risk than actually exists.

Participants emphasised the importance of targeted, age-appropriate educational approaches, particularly for children, to promote awareness without fostering fear. They also expressed interest in future research examining the effectiveness of different communication methods, including signage and engagement with community groups.

### Fieldwork context

Stakeholders described the practical challenges of conducting tick sampling and noted that interactions with the public were common during field activities, providing opportunities to offer preventive advice.

## Discussion

Insights from stakeholder focus groups highlighted key considerations for designing effective public health messaging on ticks in urban green spaces. Public awareness is inconsistent, shaped by prior exposure to rural environments, local knowledge, and personal experiences. This variability underscores the importance of identifying priority groups who may have lower awareness or hold misconceptions.

Misinformation, especially concerning tick removal, remains prevalent, indicating a need for consistent, evidence-based messaging. Participants’ observations of variable clinical advice suggest that healthcare professionals may also represent an important audience for targeted information.

Mixed views on signage suggest that communication strategies must balance clarity and reassurance. Messages that overstate risk may cause unnecessary anxiety, whereas minimal communication may leave gaps in understanding. Importantly, effective risk communication should not only highlight the potential consequences of tick bites but also provide clear and achievable prevention behaviours. This balance between communicating threat and promoting practical coping responses is consistent with Protection Motivation Theory [7] and may help increase adoption of preventive behaviours while avoiding unnecessary concern. Education targeted at children was viewed as particularly important, offering opportunities to instil preventive behaviours early without discouraging engagement with nature.

Priorities include addressing misinformation, improving consistency of advice and evaluating signage.

## Funding

This study received no external funding.

## Data availability

Anonymised thematic summaries are available from the corresponding author upon reasonable request.

## Supplementary materials

**Table S1.** Themes, sub-themes, and illustrative quotations.

| Theme | Sub-theme | Illustrative quotation |
|---|---|---|
| **Public awareness, knowledge and preventive behaviours** | Awareness of tick-borne disease | "They knew… Lyme disease was a thing, but they didn’t seem to know an awful lot about the symptoms or the occurrence of it." |
|  | General public knowledge of ticks | “I think it varies tremendously. You'll get the occasional person that I would say is very tick…tick aware they'll have the trousers tucked into the socks and if you chat to them they'll say ohh yes, when I get home, I'm going to check that I haven't picked up any ticks. And then you get the other type of person who's probably wearing sandals and shorts.” |
|  | Misinformation & incorrect procedures | “My friend says use Vaseline or. Ohh, I saw a YouTube video about how you have to twist it to the left and then the right 3 1/2 times and then do a loop the loop and you guys got there. Just get a tick tool and just pull it off.” |
|  | Importance of local knowledge | “We also try and make an effort to speak to people in that area because what we found is that local people have got a really good knowledge.” |
|  | GP knowledge variability | “Do you think it is a bit of a lottery in terms of GP, there was to whether they'll be test you was kind of a difficult issue, but some people just won't give you antibiotics” |
| **Public perceptions and responses to tick risk** | Fear or panic about ticks | “ohh, it sucks your blood, you know like feeds on the blood that that really freaks people out. And then you tell them about, you know, they also carry diseases. And that completely freaked them out.” |
|  | Personal experiences shared | “I get them on my dog or I've had them here or they'll say what's the tip. But I've had Lyme disease. Yeah…or my friend or they'll volunteer quite quickly as soon as we say what we're doing” |
|  | Acceptance of ticks outdoors | “So the staff had kind of accepted that ticks were a part of life living there and they were quite happy to just remove them that advise the guests about the risks and just remove them from them as needed” |
| **Communication & Education** | Signage and written information | “To warn people or to tell, to tell people to stay on the path or tell people to do their socks over their trousers or warn people. But I'd be quite interested in it. How much signage works.” |
|  | Questions for future research | “Educate children. Not in a way to scare them out of the countryside.<br>Combine with a science experiment. Tricky ticks classroom toolkit<br>Tick check on Paddington bear. Learning about them. Bug hunts.” |
|  | Practical advice | “They specifically ask me to say ohh well, what do we how do we stop them biting? I just say things like I'll just, you know, usual put your socks over your trousers as well…long sleeves, stay on the path.” |
| **Fieldwork Context & Practices** | Fieldwork timing & vegetation | “We tend to do a load all of our field work from kind of April, may, June time. We wouldn't really like to be doing field work, we like to look at density assessments. So you need good numbers of ticks and good activity and low vegetation levels and yeah. So by now the brambles are too high, the nettles are too high, the Bracken's too high.” |

## Notes

### Competing Interest Statement

The authors have declared no competing interest.

## References

1. Rochlin I, Toledo A. Emerging tick-borne pathogens of public health importance: a mini-review. J Med Microbiol. 2020;69(6):781

2. Gandy SL, Hansford KM, Medlock JM. Possible expansion of Ixodes ricinus in the United Kingdom identified through the Tick Surveillance Scheme between 2013 and 2020. Med Vet Entomol. 2023;37(1):96–104.

3. Collini M, Albonico F, Rosà R, Tagliapietra V, Arnoldi D, Conterno L, Rossi C, Mortarino M, Rizzoli A, Hauffe HC. Identification of Ixodes ricinus blood meals using an automated protocol with high-resolution melting analysis (HRMA) reveals the importance of domestic dogs as larval tick hosts in Italian alpine forests. Parasites Vectors. 2016;9:1–10.

4. Hansford KM, Fonville M, Gillingham EL, Coipan EC, Pietzsch ME, Krawczyk AI, Vaux AG, Cull B, Sprong H, Medlock JM. Ticks and Borrelia in urban and peri-urban green space habitats in a city in southern England. Ticks Tick Borne Dis. 2017;8(3):353–61.

5. Oliver K, Lorenc T, Innvær S. New directions in evidence-based policy research: a critical analysis of the literature. Health Research Policy and Systems. 2014;12:34.

6. Gale NK, Heath G, Cameron E, Rashid S, Redwood S. Using the framework method for the analysis of qualitative data in multi-disciplinary health research. BMC Med Res Methodol. 2013;13:117.

7. Milne S, Sheeran P, Orbell S. Prediction and intervention in health-related behavior: A meta-analytic review of Protection Motivation Theory. Journal of Applied Social Psychology. 2000;30(1):106–143.

